# MELD-XI Predicts Severe Right Ventricular Failure after HeartMate 3 Implantation in a Contemporary Cohort

**DOI:** 10.1101/2024.07.09.24310179

**Authors:** David S. Lambert, Ana María Picó, Justin D. Vincent, Elena Deych, Erin Coglianese, Joel D. Schilling, Justin M. Vader, Bin Q. Yang

## Abstract

**Background:** Right ventricular failure (RVF) after left ventricular assist devices (LVAD) is associated with significant morbidity and mortality and identifying patients at risk for severe RVF is an important clinical goal. Current risk prediction models were not developed in contemporary LVAD populations and have limited clinical applicability.

**Objectives:** To evaluate whether the Model for End Stage Liver Disease – eXcluding INR (MELD-XI) can predict severe RVF after HeartMate 3 (HM3) implantation.

**Methods:** We retrospectively analyzed all adult patients who received HM3 LVAD as initial implantation at two academic medical centers. We assessed whether MELD-XI is an independent risk factor for severe RVF in multivariate analysis and compared the predictive accuracy of MELD-XI with previously published risk scores. We also investigated the relationship between MELD-XI and markers of right ventricular function and whether MELD-XI was associated with death or pump exchange at 1-year follow-up.

**Results:** Our study included a total of 246 patients, of which 74 (30%) experienced severe RVF. After adjusting for relevant covariables, MELD-XI was independently associated with severe RVF (OR 1.18, CI 1.09-1.29, p<0.001) and performed similarly to the EUROMACS and Michigan RVF risk scores. In addition, MELD-XI was not reflective of traditional echocardiographic or hemodynamic measures of right ventricular function. Finally, MELD-XI ≥ 14 predicted worse in-hospital mortality.

**Conclusions:** Among patients undergoing HM3 implantation, MELD-XI is independently associated with an increased risk of RVF and in-hospital mortality.

## Introduction

Left ventricular assist devices (LVAD) are indicated for the treatment of end-stage heart failure (HF). Improvements in LVAD technology, specifically the utilization of the HeartMate 3 (HM3) continuous flow device, have significantly improved clinical outcomes.^1^ However, post-LVAD right ventricular failure (RVF) remains a frequent complication that leads to an increased risk of renal failure, stroke, prolonged hospitalization, and death.^2–5^ Given the high morbidity and mortality associated with RVF, multiple scores have been developed to estimate the risk of severe RVF following LVAD implantation.^5–9^ However, these scores were developed in predominantly non-HM3 populations and external validation attempts have shown reduced predictive accuracy.^10–11^ Thus, an easy-to-use and accurate method to predict severe RVF in the contemporary era remains an unmet need in the field.

The liver is the first organ upstream of the right-sided heart chambers and is sensitive to elevated central venous pressures. Congestive hepatopathy is a recognized complication of heart failure that can lead to hepatic dysfunction and in some cases cardiac cirrhosis.^12–13^ The Model for End Stage Liver Disease – eXcluding INR (MELD-XI) is a modification of the original MELD that was created for use in patients requiring anticoagulation, which is common among those with HF.^14^ Elevated MELD-XI has been associated with worse survival in hospitalized HF patients and LVAD recipients and may predict RVF in a non-HM3 population.^15–17^ Whether MELD-XI is an independent risk factor for severe RVF in HM3 recipients has not been explored.

Therefore, in this multicenter study, we sought to investigate whether MELD-XI is associated with an increased risk of severe RVF in patients undergoing HM3 implantation and to compare the performance of MELD-XI with previously published risk scores. We hypothesized that higher MELD-XI would be an independent risk factor for severe RVF and 1-year mortality.

## Methods

### Study population

We retrospective analyzed all adult patients over the age of 18 who received a HM3 LVAD as initial implantation between 2015 and 2022 at Washington University in St. Louis and Massachusetts General Hospital. We excluded patients who underwent pump exchange, had insufficient clinical data, or less than 1 year of follow-up. Patient demographics and comorbidities were extrapolated from institutional LVAD databases. Laboratory values were taken immediately prior to surgery and echocardiographic and hemodynamic data were obtained from the most recent transthoracic echocardiogram or right heart catheterization within 90 days, respectively. This study was approved by the IRBs at Washington University in St. Louis (202205108) and Massachusetts General Hospital (2018P000131).

### MELD-XI and RVF risk scores

MELD-XI, a previously validated measure of liver dysfunction developed for patients who may be on anticoagulation, was calculated as 5.11*Ln(bilirubin) + 11.76*Ln(creatinine) + 9.44.^14^ The EUROMACS score was calculated as right atrial pressure to pulmonary capillary wedge pressure ratio (RAP/PCWP) > 0.54 (2 points) + hemoglobin ≤ 10g/dL (1 point) + multiple intravenous inotropes (2.5 points) + INTERMACS profile 1-3 (2 points) + severe RV dysfunction on echocardiography (2 points).^8^ The Michigan RVF risk score was calculated as vasopressor requirement (4 points) + AST ≥ 80 IU/l (2 points) + bilirubin ≥ 2.0 mg/dl (2.5 points) + creatinine ≥ 2.3 mg/dl (3 points).^9^

### Outcomes

The primary outcome was severe RVF according to the INTERMACS definition, characterized by clinical symptoms of elevated filling pressures and needing right ventricular assist device implantation, > 14 days of inotropes, or > 48 hours of pulmonary vasodilators following HM3 implantation.^2^ Secondary outcomes were in-hospital death and a composite of post-discharge death or pump exchange. All patients were followed for at least 1 year and censored at the last date of follow-up or heart transplant, the latter of which was considered a successful endpoint.

### Statistical analysis

Clinical and demographic variables were compared between subjects with and without severe RVF. Continuous variables are presented as mean (STD) or median (IQR) and compared using a t-test or Wilcoxon test, depending on the distribution. Categorical and ordinal variables are compared using the Chi-squared and Cochran-Armitage trend tests, respectively. MELD-XI, EUROMACS, and Michigan risk scores were individually calculated for each subject, and receiver operator curves are shown with their AUC for predicting severe RVF. The correlation between MELD-XI and echocardiographic and hemodynamic measures of RV dysfunction was assessed visually and by the Spearman correlation coefficient. To evaluate the effect of MELD-XI on severe RVF, we fit a generalized linear mixed effects model with a random study center effect, using binomial distribution. The variables offered into the model were age, sex, race, NT-proBNP, RAP/PCWP ratio, pulmonary artery pulsatility index (PAPi), and INTERMACS profile. The final model was selected by backward elimination using AIC criteria. Survival was estimated using Kaplan-Meier curves with log-rank test, stratified *a priori* by MELD-XI ≥ or < 14, and compared using log-rank test. In addition, multivariable Cox proportional hazards models were fitted with random study center effect and separately for in-hospital and post-discharge outcomes of mortality or pump exchange. Proportionality assumptions were tested for all Cox models. All analyses were performed in R, version 4.3.1.

## Results

### Baseline characteristics

Our study included 246 HM3 recipients, of which 28% were African-American/black and 20% were female. In total, 74 (30%) of patients experienced severe RVF by the INTERMACS definition. When comparing those who developed severe RVF to those who did not, baseline demographics and medical comorbidities were similar. However, at time of implantation, patients who developed severe RVF were more likely to be INTERMACS profile 1 or 2 (50% vs. 26%, p<0.001), have a higher RAP/PCWP (0.58 vs. 0.42, p <0.001), lower PAPi (1.5 vs. 2.5, p<0.001), and elevated MELD-XI (16.56 vs. 13.21, *p*<0.001, **Table 1**). Between the two institutions, patient demographics were similar but those implanted at Washington University in St. Louis had more high-risk features, with a greater percentage of INTERMACS profile 1 or 2 (42.2% vs. 13.8%), higher RAP/PCWP (0.53 vs. 0.36), and higher MELD-XI (14.54 vs. 13.18, Supplemental table 1).

**Table 1:** Baseline characteristics of patients undergoing HeartMate 3 implantation. RVF=right ventricular failure, HF=heart failure, LVEF=left ventricular ejection fraction, RAP/PCWP=right atrial pressure/pulmonary capillary wedge pressure, TAPSE=tricuspid annular plane systolic excursion, PAPi=pulmonary artery pulsatility index. All continuous variables are expressed as median [IQR].

|  | Overall | Non-RVF | Severe RVF | p |
| --- | --- | --- | --- | --- |
| n | 246 | 172 | 74 |  |
| Age (mean (SD)) | 58 (13) | 59 (12) | 56 (14) | 0.118 |
| Sex =<br>Female/Male (%) | 49/197 (20/80) | 31/141 (18/82) | 18/56 (24/76) | 0.337 |
| Race = Non-White/White (%) | 68/177 (28/72) | 46/126 (27/73) | 22/51 (30/70) | 0.699 |
| HF etiology<br>Ischemic/Non-<br>ischemic (%) | 102/143 (42/58) | 74/97 (43/57) | 28/46 (38/62) | 0.515 |
| INTERMACS<br>profile (%) |  |  |  | <0.001 |
| 1-2 | 81 (33) | 44 (26) | 37 (50) |  |
| 3 | 118 (48) | 87 (51) | 31 (42) |  |
| 4-7 | 47 (19) | 41 (24) | 6 (8) |  |
| LVEF | 18 [14, 22] | 18 [14, 23] | 18 [13, 20] | 0.139 |
| RAP/PCWP | 0.47 [0.31, 0.66] | 0.42 [0.29, 0.59] | 0.58 [0.44, 0.77] | <0.001 |
| TAPSE | 1.60 [1.30, 1.90] | 1.60 [1.40, 2.00] | 1.50 [1.28, 1.80] | 0.098 |
| PAPi | 2.00 [1.30, 3.60] | 2.50 [1.60, 4.04] | 1.50 [1.10, 2.44] | <0.001 |
| Log(NT-proBNP) | 8.55 [7.96, 9.17] | 8.53 [7.86, 9.13] | 8.56 [8.15, 9.44] | 0.221 |
| Cardiac output | 3.73 [3.00, 4.50] | 3.70 [3.00, 4.50] | 3.85 [3.00, 4.30] | 0.750 |
| Bilirubin | 0.90 [0.52, 1.40] | 0.80 [0.50, 1.20] | 1.20 [0.70, 2.08] | <0.001 |
| Creatinine | 1.33 [1.05, 1.75] | 1.27 [1.01, 1.68] | 1.44 [1.17, 1.94] | 0.004 |
| MELD-XI | 14 [11, 17] | 13 [11, 16] | 17 [12, 19] | <0.001 |

### MELD-XI is an independent risk factor for severe RVF after HM3 implantation

We next analyzed whether MELD-XI is an independent risk factor for severe RVF. Our initial model adjusted for age, sex, race, NT-proBNP, INTERMACS profile, RAP/PCWP, PAPi, and MELD-XI. After backward elimination using AIC criteria, age, sex, race, NT-proBNP, and PAPi were all found to be nonsignificant in a stepwise fashion and thus our final model incorporated only INTERMACS profile, RAP/PCWP, and MELD-XI to optimize performance in predicting the primary outcome. We found that MELD-XI was independently associated with severe RVF, with each 1-point increase in MELD-XI corresponding to an 18% increased risk of severe RVF (p<0.001, **Table 2**). In addition, RAP/PCWP (OR 1.33 for every 0.1-point increase in RAP/PCWP ratio, p<0.001) and INTERMACS profile 1 or 2, when compared to 4 (OR 3.11, p=0.032), were risk factors for severe RVF as well.

**Table 2:** MELD-XI is an independent predictor of severe right ventricular failure (RVF) after HeartMate 3 implantation. Our initial mode adjusted for age, sex, race, NT-proBNP, INTERMACS profile, right atrial pressure/pulmonary capillary wedge pressure (RAP/PCWP), pulmonary artery pulsatility index (PAPi), and MELD-XI. After backwards elimination of nonsignificant variables age, race, sex, NT-proBNP, and PAPi, our final model incorporated MELD-XI, RAP/PCWP, and INTERMACS profile. ^∗^OR per 1 point increase in MELD-XI score. ^†^OR per 0.1 increase in RAP/PCWP ratio. ^‡^OR compared to INTERMACS 4-7.

| Variable | OR | 95% CI | p |
| --- | --- | --- | --- |
| MELD-XI* | 1.18 | 1.09-1.29 | <0.001 |
| RAP/PCWP† | 1.33 | 1.14-1.54 | <0.001 |
| INTERMACS profile 1-2‡ | 3.12 | 1.10-8.84 | 0.032 |
| INTERMACS profile 3‡ | 1.58 | 0.57-4.36 | 0.377 |

### MELD-XI performs as well as previously published risk scores

Since MELD-XI was associated with the risk of severe RVF in our patient population, we sought to compare the predictive accuracy of MELD-XI to previously published risk prediction tools.

Two of the most commonly used RVF risk scores are the EUROMACS, which is the most contemporary and only model that included HM3 recipients, and the Michigan RVF score, which is the most widely validated to date.^11^ We first validated the EUROMACS (AUC 0.68) and Michigan RVF score (AUC 0.66) in predicting severe RVF in our cohort and subsequently found that MELD-XI performed equally (AUC 0.69, **Figure 1**).

**Figure 1:**
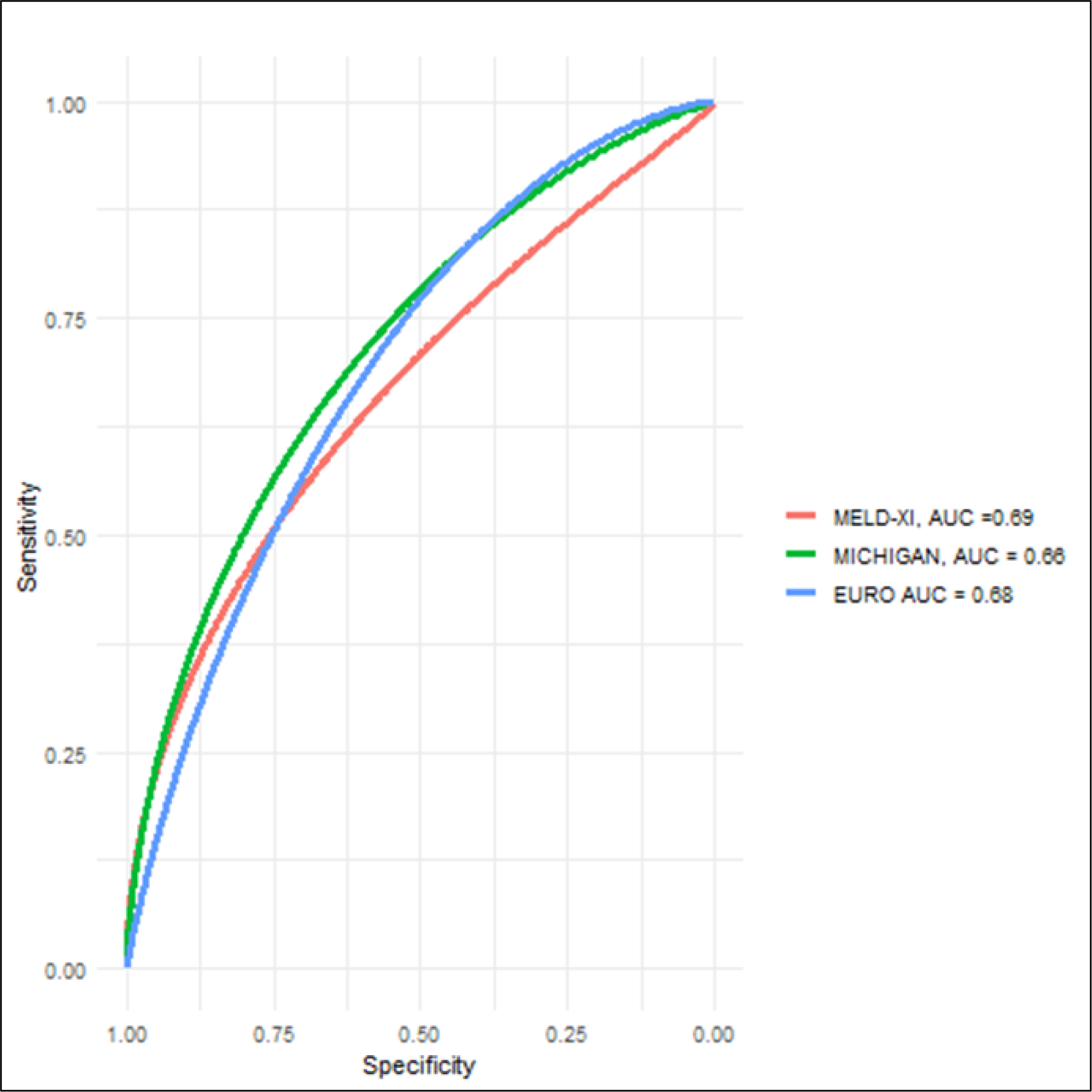
MELD-XI performs similarly to previously published risk scores to predict severe right ventricular failure (RVF) after left ventricular assist device implantation. Comparison of AUC between EUROMACS (black), Michigan RVF (blue), and MELD-XI (red) in this multi-institutional cohort of HeartMate 3 recipients.

### MELD-XI does not correlate with measures of RV function

As MELD-XI is a surrogate marker of hepatorenal disease which could reflect chronic congestion in patients with HF, we next asked whether it correlated with echocardiographic or hemodynamic measures of RV dysfunction.^2,11, 18–24^ Frequently used parameters include tricuspid annular plane systolic excursion (TAPSE), RAP/PCWP ratio, and PAPi. Interestingly, we found that MELD-XI did not or poorly correlated with all of the following variables: TAPSE (r = - 0.15), RAP (r = 0.22), RAP/PCWP ratio (r = 0.16), mean pulmonary artery pressure (r = 0.27), PAPi (r = -0.09), and pulmonary vascular resistance (r = 0.07, **Figure 2**).

**Figure 2:**
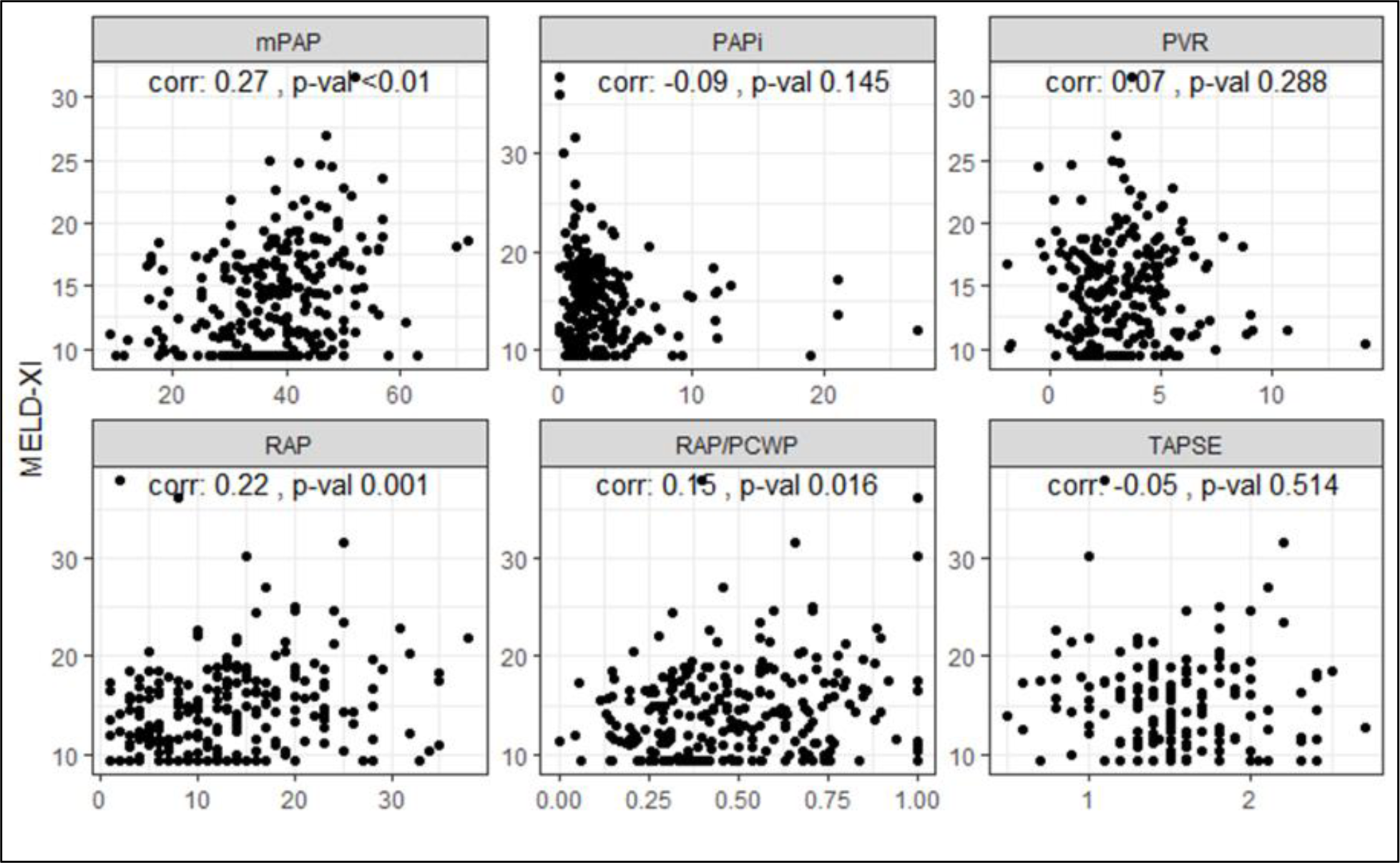
MELD-XI does not or poorly correlate with echocardiographic and hemodynamic measures of right ventricular (RV) function. **Dot plots of the relationship** between MELD-XI and different measures of RV function and pulmonary artery hemodynamics. Each dot represents an individual patient. mPAP=mean pulmonary artery pressure, PAPi=pulmonary artery pulsatility index, PVR=pulmonary vascular resistance, RAP=right atrial pressure, PCWP=pulmonary capillary wedge pressure, TAPSE= tricuspid annular planar systolic excursion.

### MELD-XI predicts early but not late mortality after HM3 implantation

Finally, we assessed whether preoperative MELD-XI can predict mortality after HM3 implantation. We stratified patients by MELD-XI ≥ 14 or < 14 *a priori* and found that MELD-XI ≥ 14 was associated with significantly worse in-hospital mortality after LVAD implantation (log-rank p=0.002). However, MELD-XI did not predict the risk of death or pump exchange at 1-year follow-up among patients who survived index hospitalization (log-rank p = 0.880, **Figure 3**).

**Figure 3:**
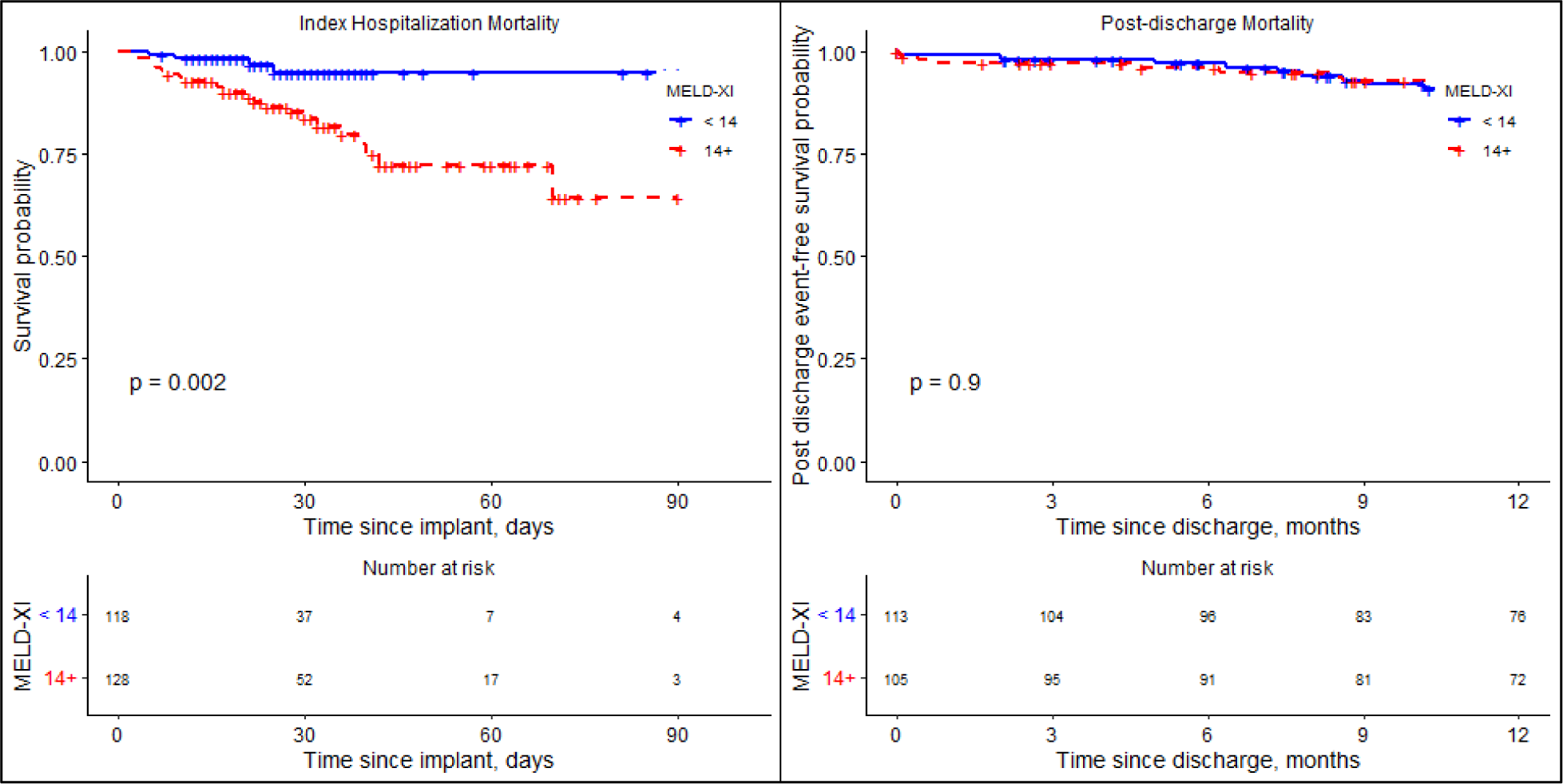
MELD-XI predicts early but not late mortality or pump exchange after **HeartMate 3 implantation.** Kaplan-Meier curves are shown, stratified by MELD-XI ≥ 14 (red) and < 14 (blue). Patients with higher MELD-XI had worse in-hospital mortality (left, log rank p=0.002), but MELD-XI did not predict survival after discharge from index hospitalization (right, log rank p=0.9).

The effect of MELD-XI was confirmed in the multivariate Cox proportional hazard model. After adjusting for age and RAP/PCWP, higher MELD-XI was associated with increased mortality during index hospitalization (HR 1.10 per 1-point increase, CI 1.03-1.18, p < 0.01) but not after discharge (HR 1.02, CI 0.92-1.14 p=0.679, **Supplemental Table 2**). Lastly, given the changing demographics of LVAD recipients after the 2018 UNOS heart transplant allocation system change, we analyzed whether MELD-XI still performs well in the contemporary era.^25–26^ We found that MELD-XI ≥ 14 consistently predicted early but not late mortality before and after the date of the allocation system change (**Figure 4**).

**Figure 4:**
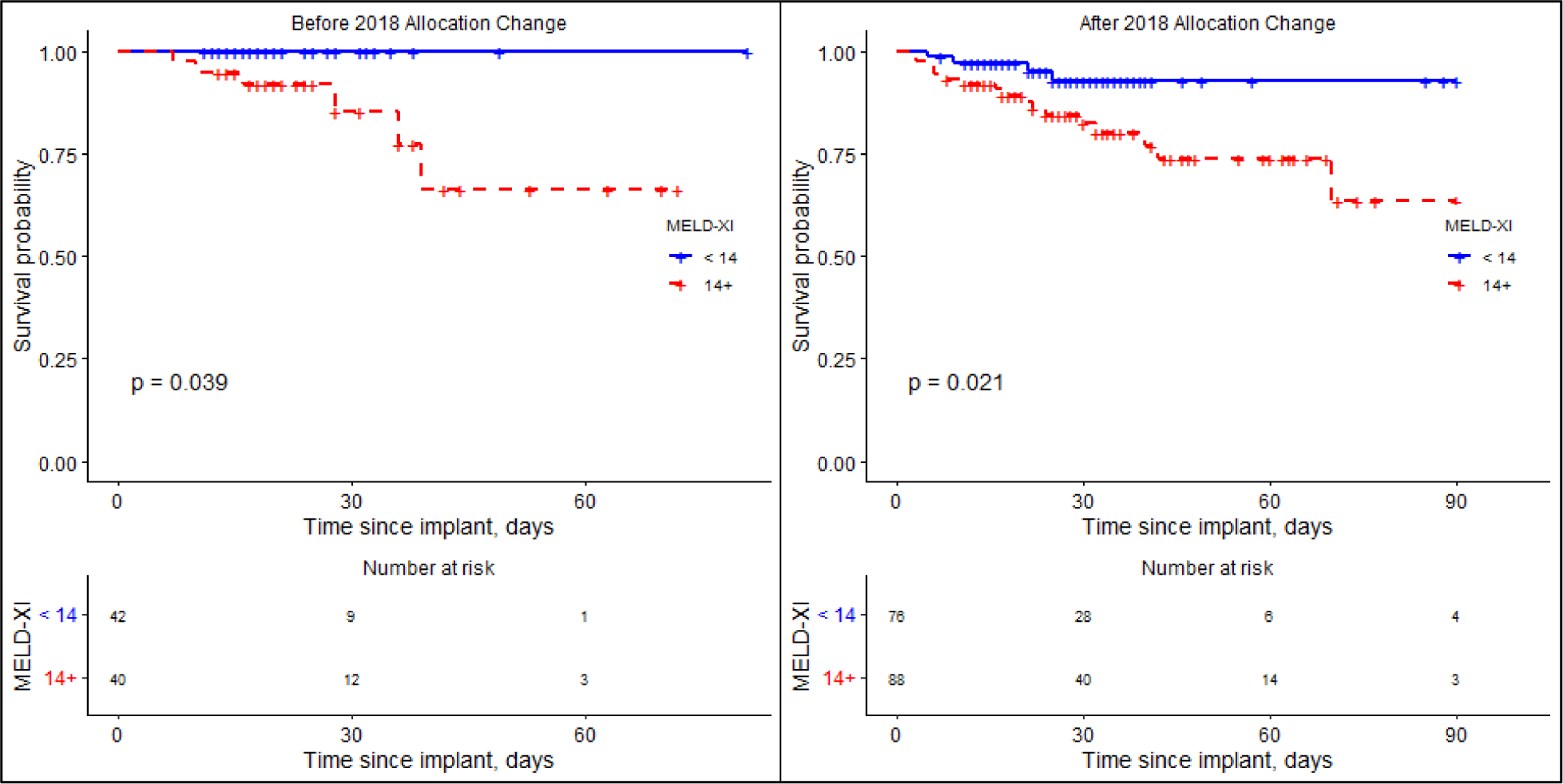
MELD-XI remains predictive of in-hospital mortality in the contemporary era. Kaplan-Meier curves are shown before (left) and after (right) the 2018 heart transplant allocation system change. Patients with MELD-XI ≥ 14 (red) had higher in-hospital mortality or pump exchange than those with MELD-XI < 14 (blue) in both eras (log rank p=0.039 and 0.021 for before and after, respectively).

## Discussion

In this multicenter retrospective analysis, we found that MELD-XI was independently associated with severe RVF and in-hospital mortality after HM3 implantation. In addition, MELD-XI performed just as well as previously validated RVF risk scores but interestingly, did not correlate with traditional echocardiographic and hemodynamic measures used to assess RV function.

These findings shed important insights into predictors of clinical outcomes in contemporary LVAD recipients and the syndrome of RVF.

### RVF is the major cause of early mortality and previous risk scores have limited applicability in the contemporary era

The occurrence of severe RVF after LVAD is a major driver of in-hospital mortality as well as significant morbidity including the need for renal replacement therapy, stroke, and gastrointestinal bleeding.^2–4, 27–29^ Thus, identification of patients at increased risk of RVF is paramount in the evaluation of patients for LVAD candidacy, in shared decision-making to proceed with HM3 implantation, and perioperative planning. Previous studies have analyzed preoperative risk factors that increase the likelihood of severe RVF, using a combination of hemodynamic, echocardiographic, and clinical variables to generate a weighted risk score. Of all the scores, EUROMACS is the only one to include HM3 recipients; however, patients with this device comprised only 9% of the derivation cohort.^8^ The Michigan RVF risk score, on the other hand, is the most validated tool to date but was developed during a time when only 14% of patients had continuous flow LVAD devices.^9,11^ Furthermore, multiple external validation attempts have yielded disappointing results.^10,30^ Therefore, identifying an easy-to-use risk prediction tool applicable to HM3 recipients remains an unmet clinical need.

### MELD-XI is simple, widely available, and has broad application

MELD-XI was originally introduced by Heuman et al to account for the high prevalence of anticoagulation use in HF patients, which correlated well with MELD in cirrhotic patients.^14^ Elevated MELD and MELD-XI have been shown to be associated with a higher risk of mortality in patients undergoing LVAD.^17,31–32^ However, these studies were single-centered and conducted in an era of non-HM3 devices. In the present study, we show that higher MELD-XI is associated with an increased likelihood of severe RVF and the discriminatory power for severe RVF is comparable to previous risk scores, many of which require 4 or more variables. On the other hand, MELD-XI is easy to calculate and only relies on bilirubin and creatinine, two readily available clinical lab values. Moreover, we found that MELD-XI predicted in-hospital mortality and was not impacted by the UNOS allocation system change in October 2018, making it a powerful tool for preoperative risk assessment in patients undergoing evaluation for LVAD in the contemporary era. In contrast to previous studies, we also found that MELD-XI was not associated with mortality beyond index hospitalization. First, this further supports our primary finding that MELD-XI is a robust predictor of severe RVF, the key driver of early mortality.

Second, hemodynamic support by HM3 may lead to regression of hepatorenal disease over time if patients survive the initial postoperative period. This is in line with Yang et al, who showed that patients who experienced an improvement in MELD-XI post-LVAD had similar survival after heart transplant to those who had a low MELD-XI to begin with.^33^ More recently, Mehra et al derived and validated the HM3 risk score (HM3RS) in the Multicenter Study of MagLev Technology in Patients Undergoing Mechanical Circulatory Support Therapy with HeartMate 3 (MOMENTUM 3) clinical trial population.^34^ The HM3RS also incorporates measures of both renal and RV dysfunction (BUN and RAP/PCWP, respectively). However, exploratory analysis in our cohort showed that HM3RS did not predict severe RVF (AUC 0.48), possibly related to its not incorporating markers of liver disease.

### Hepatorenal dysfunction and the syndrome of right heart failure

Intriguingly, we found that while MELD-XI predicted severe RVF, it does not correlate with echocardiographic or hemodynamic measures of RV function used clinically. In predominantly single-centered studies, lower TAPSE and higher PAPi and RAP/PCWP have all been shown to increase the risk of severe RVF, which is in line with our finding that RAP/PCWP (although not TAPSE/PAPi) is an independent risk factor for severe RVF. In our cohort, MELD-XI did not correlate or correlated poorly with measures of RV systolic function (TAPSE), right-sided filling pressure (RAP), pulmonary artery pressure, or relative RV performance (PAPi, RAP/PCWP).

Taken together, MELD-XI seems to provide additional and incremental information about the risk of RVF in patients undergoing LVAD implantation independent of aforementioned measures of RV function. Increasingly, RVF is recognized as a systemic syndrome characterized by end-organ dysfunction due to chronic venous congestion.^13,34–35^ More recently, our group has shown that patients with clinical RVF have a unique systemic inflammatory profile and some of these circulating factors may be released by Kupffer cells, which are resident hepatic macrophages.^36^ We hypothesize that MELD-XI reflects the global impact of RVF over time in a way that is not captured by snapshot measurements inherent in echocardiographic or hemodynamic parameters. (**Central Illustration**) The worst outcomes were observed in patients with high MELD-XI and RAP/PCWP ratio. Therefore, integrating RV dysfunction severity (RAP/PCWP ratio) and its downstream consequences (MELD-XI) may better capture the overall phenotypic profile of RVF than either alone. Exploratory analysis showed that the combination of MELD-XI and RAP/PCWP had an AUC 0.76 in predicting severe RVF and validating this finding should be an area of future investigation.

## Limitations

Our study has several limitations. First, it is a retrospective analysis of patients treated at two academic medical centers. However, this is one of the larger HM3-exclusive studies to date. In addition, the primary findings were consistent across both institutions, increasing the generalizability of our findings. Second, we acknowledge that there are multiple risk scores to predict severe RVF.^11^ We picked the EUROMACS and Michigan scores because they are the only ones to incorporate HM3 recipients and have been the most widely validated, respectively. Finally, while MELD-XI is associated with severe RVF, our study suggests that combining MELD-XI and RAP/PCWP ratio may offer even more predictive accuracy. This needs to be validated in an external cohort.

## Conclusion

MELD-XI is independently associated with an increased risk of severe RVF and in-hospital mortality after HM3 implantation.

## Data Availability

All data will be available upon request of corresponding author.

## Acknowledgements

The authors would like to thank the Mentors in Medicine program at Washington University School of Medicine in St. Louis.

## Supplemental Material

Table S1 Table S2

## Clinical Perspectives

- Despite the improving engineering design of LVAD leading to better clinical outcomes, post-LVAD RVF remains a major driver of early morbidity and mortality.
- To fully capture the phenotype and risk of RVF, clinicians should integrate measures of both RV dysfunction severity (RAP/PCWP ratio) and chronicity (MELD-XI) to help select appropriate patients for LVAD therapy and perioperative planning.
- Future studies should focus on validating the combination of RAP/PCWP and MELD-XI in predicting severe RVF and incorporating these variables into clinical decision-making.

## Non-standard Abbreviations and Acronyms

LVAD: left ventricular assist device
HF: heart failure
HM3: HeartMate 3
RVF: right ventricular failure
MELD-XI: Model for End Stage Liver Disease – eXcluding INR
RAP: right atrial pressure
PCWP: pulmonary capillary wedge pressure
PAPi: pulmonary artery pulsatility index
HM3RS: HeartMate 3 risk score
MOMENTUM 3: Multicenter Study of MagLev Technology in Patients Undergoing Mechanical Circulatory Support Therapy with HeartMate 3

---

**Central Illustration:**
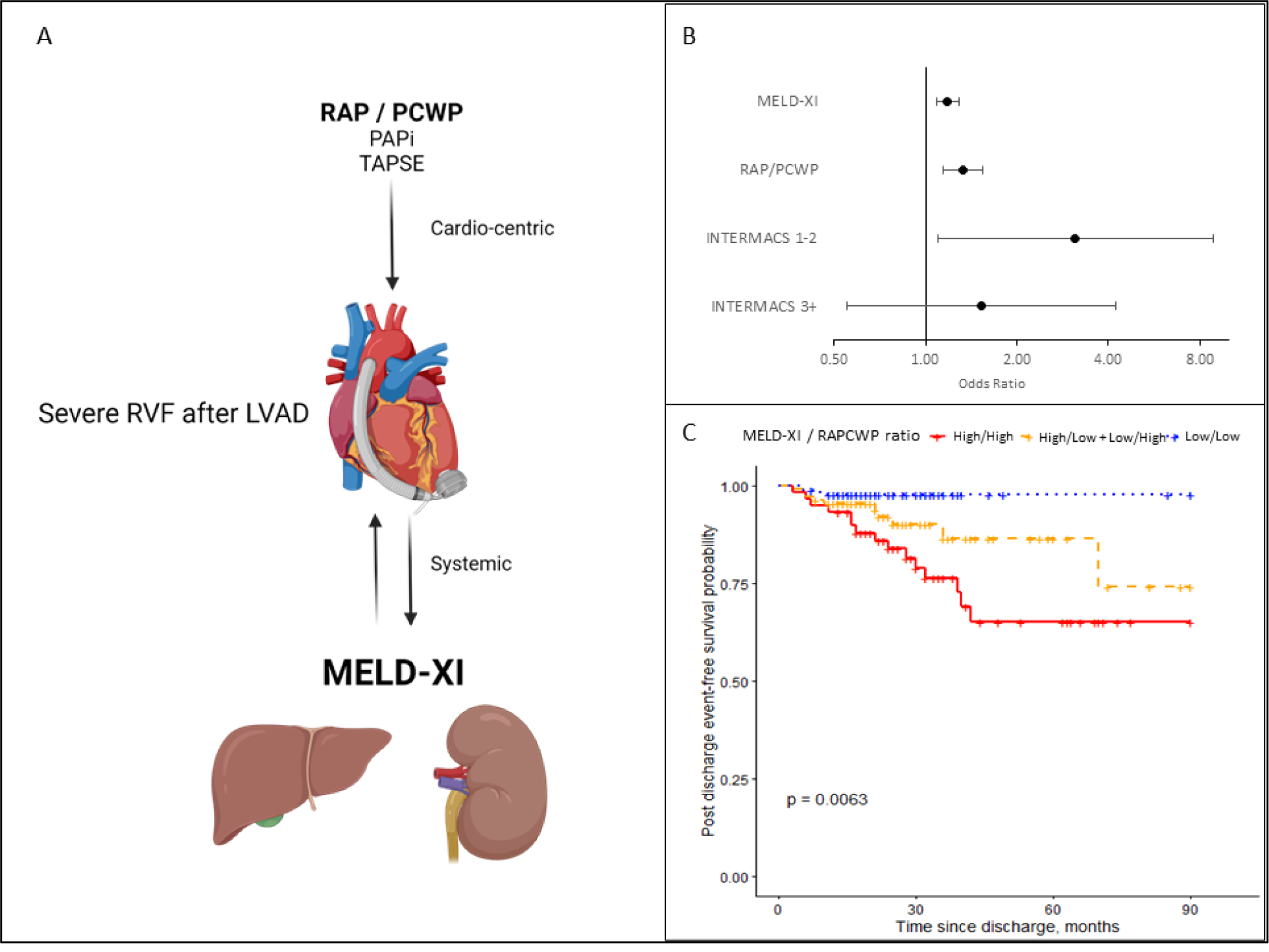
Preoperative MELD-XI predicts severe right ventricular failure (RVF) and in-hospital mortality following HeartMate 3 (HM3) implantation. **We propose** that clinicians should integrate both measures of RV function and systemic consequences of right heart failure in their assessment of preoperative risk of severe RVF after HM3 (Panel A). MELD-XI is an independent risk factor for severe RVF in this multi-institutional cohort (Panel B) and the combination of right atrial pressure/pulmonary capillary wedge pressure (RAP/PCWP) ≥ 0.55 and MELD-XI ≥ 14 portends the worst in-hospital mortality (Panel C). Created with BioRender.

